# DeePaN: A <u>dee</u>p <u>pa</u>tient graph convolutional <u>n</u>etwork integrating clinico-genomic evidence to stratify lung cancers benefiting from immunotherapy

**DOI:** 10.1101/19011437

**Authors:** Chao Fang, Dong Xu, Jing Su, Jonathan R Dry, Bolan Linghu

## Abstract

Immuno-oncology (IO) therapies have transformed the therapeutic landscape of non-small cell lung cancer (NSCLC). However, patient responses to IO are variable and influenced by a heterogeneous combination of health, immune and tumor factors. There is a pressing need to discover the distinct NSCLC subgroups that influence response. We have developed a <u>dee</u>p <u>pa</u>tient graph convolutional <u>n</u>etwork, we call “DeePaN”, to discover NSCLC complexity across data modalities impacting IO benefit. DeePaN employs high-dimensional data derived from both real world evidence (RWE) based electronic health records (EHRs) and genomics across 1,937 IO treated NSCLC patients. DeePaN demonstrated effectiveness to stratify patients into subgroups with significantly different (p-value of 2.2 × 10^−11^) overall survival of 20.35 months and 9.42 months post-IO therapy. Significant differences in IO outcome were not seen from multiple non-graph based unsupervised methods. Furthermore, we demonstrate that patient stratification from DeePaN has the potential to augment the emerging IO biomarker of tumor mutation burden (TMB). Characterization of the subgroups discovered by DeePaN indicates potential to inform IO therapeutic insight, including the enrichment of mutated KRAS and high blood monocyte count in the IO beneficial and IO non-beneficial subgroups, respectively. To the best of our knowledge, our work for the first time has proven the concept that graph based AI is feasible and can effectively integrate high-dimensional genomic and EHR data to meaningfully stratify cancer patients on distinct clinical outcomes, with potential to inform precision oncology.

## Introduction

Recently immuno-oncology (IO) therapies including checkpoint inhibitors have transformed the therapeutic landscape of non-small cell lung cancer (NSCLC)^1-3^. However, responses to IO in NSCLC are highly variable. Recent findings suggest a heterogeneous collection of genomic alterations and clinical phenotypes can influence IO response^4-6^. Thus, there is a pressing need to discover and characterize NSCLC subgroups across both clinical and genomic landscapes to advance precision immuno-oncology.

Real-world-evidence (RWE) based clinical phenotype data such as electronic health records (EHRs), which include patient exposures, lab data, diagnosis, medications, and clinical outcomes, represent a promising resource for precision oncology. EHR derived data has been used to identify patient subgroups to inform cancer therapeutics^7-12^. Distinct molecular subtypes^13-18^ derived from rich genomic resources, including high tumor mutational burden (TMB) and high PDL1 protein expression, have also been associated with beneficial responses to checkpoint inhibitor therapies in NSCLC^1,19-21^. The integration of both genomic and EHR evidence is expected to reveal a fuller description of tumor and patient characteristics impacting drug response. Whilst there have been many comparative studies between these high dimensional data modalities^22-24^, few studies to date integrate both genomics and EHRs for patient stratification due to all types of challenges. For instance, the study cohort can be too small to investigate this heterogeneous disease; the datasets used in subtyping studies may not be comprehensive enough to incorporate both genomic data and diverse clinical-phenotype data with long-term follow-ups; and the subtyping algorithms and models may not be effective enough to integrate high-dimensional data from both genomic and clinical domains.

Recently artificial intelligence (AI) and deep learning methods have demonstrated great potential for discovery of cancer subtypes^25-28^, stemming from effective high-dimensional data integration and capture of complex nonlinear relationships^29-31^. However, most AI studies use a grid-based model^28,32,33^ for patient data representation which overlook patient-patient relationships and are sub-optimal for inclusion of multiple data modalities. Graph based patient similarity networks (PSNs) have shown promise for patient subtyping^34,35^. PSNs effectively model patient-patient relationships to intuitively enable heterogeneous data integration and to cluster patients into subtypes based on their feature similarities. Addition of deep convolutional neural networks (CNNs) based learning of patient-data embeddings to the PSN framework holds great potential to augment patient subtype discovery through integrative usage of both genomic and EHR data.

Graph convolutional networks (GCN)^36^ are such an efficient variant of CNNs operated on a network (i.e. graph) like PSN’s. GCNs offer fast and scalable classification of nodes in a graph through graph embedding and convolutional operations. GCN has demonstrated promise in multiple biomedical applications such as protein interface prediction and side effects prediction^37^. We sought to explore a novel application of GCN on its feasibility and effectiveness for patient subtype discovery through integrative usage of EHR and genomic data.

We developed a data-driven, unsupervised, graph based AI representation we call “DeePaN” (i.e. <u>dee</u>p <u>pa</u>tient graph convolutional <u>n</u>etwork) to stratify NSCLC patients,integrating 100 EHR and genomic data features from the Flatiron Health and Foundation Medicine NSCLC “clinico-genomic” database^38^ across a cohort of 1,937 IO treated NSCLC patients. Our “DeePaN” framework employs a graph convolutional network autoencoder to learn a patient-similarity-graph based feature representation, followed by graph spectral clustering for patient subgrouping.

The “DeePaN” framework stratified patients into subgroups with distinct outcomes post-IO therapy, and this stratification was most significant when both genomic and EHR data modalities were integrated. Median survival was 9.42 months from sub-groups with poor survival vs. 20.35 months for the subgroup with better survival (p-value of 2.2× 10^−11^). Comparatively, patient sub-groupings derived through well-established methods such as autoencoder, uniform manifold approximation and projection (UMAP), and t-Distributed Stochastic Neighbor Embedding (t-SNE), showed no significant difference on IO therapy outcome. Furthermore, we demonstrated the potential to use this DeePaN grouping to augment the clinical utility of an emerging IO biomarker, tumor mutation burden (TMB). Characterization of the subgroups discovered by DeePaN indicates potential to inform IO therapeutic insight, including the enrichment of KRAS mutations and high blood monocyte count in the IO beneficial subgroup and IO non-beneficial subgroup, respectively.

“DeePaN” represents a novel graph-based AI framework with advances of effectively integrating heterogeneous clinico-genomic data modalities, leveraging graph embedding to intuitively model patient-patient relationships, and incorporating the high-performance of AI to capture nonlinear and complex relationships of patient data. To the best of our knowledge, this is the first study to demonstrate the feasibility and effectiveness of employing a graph-based AI approach to integrate RWE based high-dimensional EHRs and genomics to stratify NSCLC patients by IO benefit. The new subtypes discovered in this work may cast new light on understanding the heterogeneity of IO treatment responses, and pave ways to inform clinical decision making and therapeutics insight for precision oncology.

## Results

### Identification of an IO treated NSCLC cohort with linked clinico-genomic data

The aim of this study is to explore the feasibility and effectiveness to develop a data-driven, unsupervised, graph AI based “Deep patient graph” (DeePaN) framework integrating genomics and EHRs to stratify NSCLC patients into subgroups useful for precision immunotherapy. Using Flatiron NSCLC clinic-genomic database, we identified an IO treated cohort of 1,937 patients characterized by 100 clinico-genomic features to develop and test this framework (Methods and Figure 1). The cohort’ overall clinical and demographic characteristics are shown in Table 1 and tumor genomic characteristics are shown at Supplemental Figure 1C in Supplemental material I.

**Table 1.**
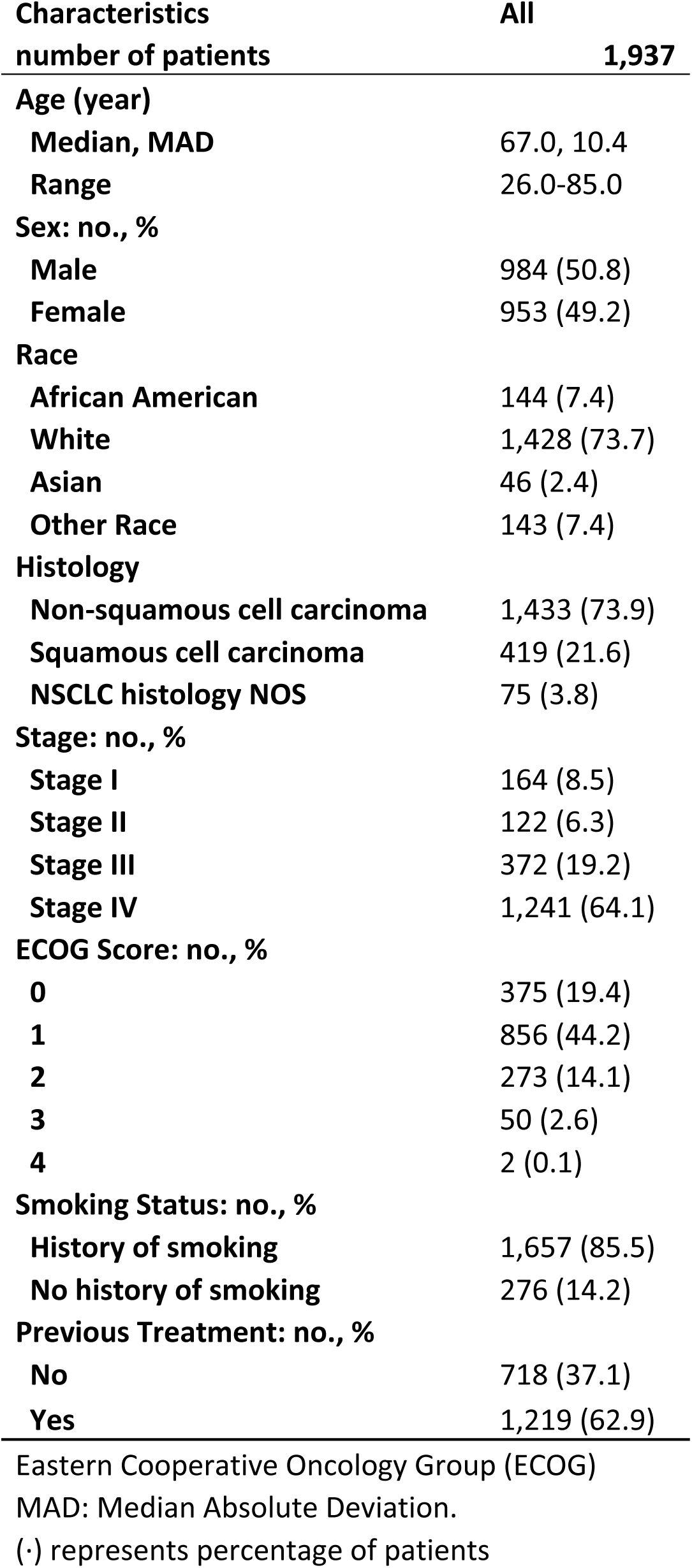
Baseline demographic and pathologic characteristics

**Figure 1.**
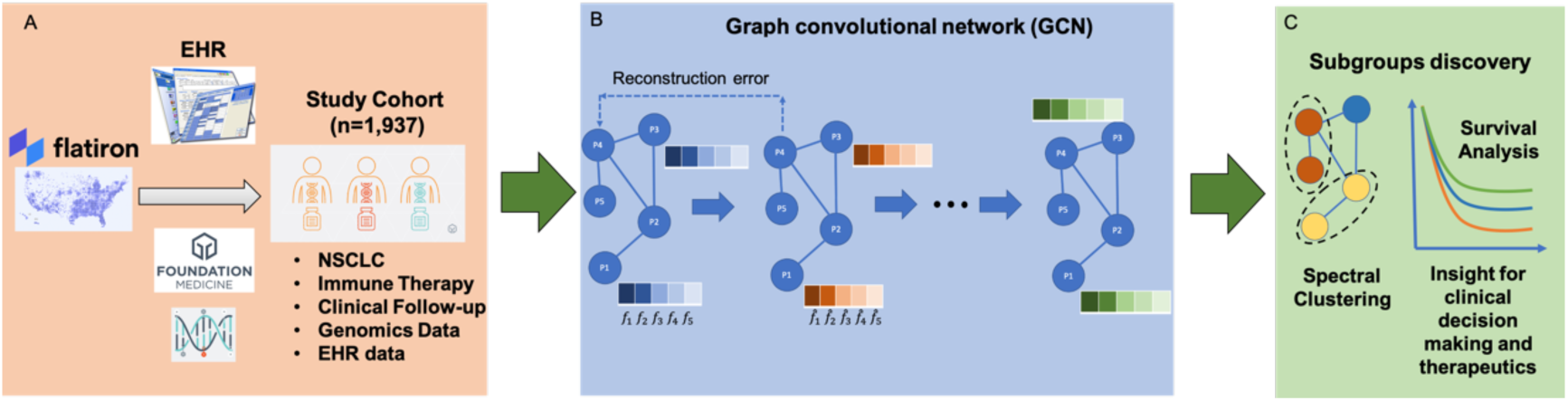
The conceptual “DeePaN” framework as a deep patient graph convolutional network integrating electronic health records and genomics to strategy NSCLC patients benefiting from immunotherapy. **A)** An IO treated NSCLC cohort (N=1,937) was identified from Flatiron clinico-genomic database with linked EHRs and genomics data. The clinical and genomic features are preprocessed (See Material and Method part for details) and concatenated as raw patient-data representations. **B)** The raw patient-data representations are modeled by a deep patient graph convolutional network (GCN) implemented as the Marginalized graph autoencoder (MGAE) to learn latent patient representations. In GCN modeling, patients are represented as nodes, and patients with similar clinico-genomic features are linked by edges. Multiple layers of graph convolutional network are stacked to learn latent patient representations, with each layer of the graph neural network being trained to produce a high-level patient data representation from the output of the previous layer. **C)** The graph-based deep patient representations are then subjective to spectral clustering to discover patient subgroups with distinct immunotherapy outcomes to inform precision-oncology including patient stratification by IO benefit.

### Construction of a deep patient graph convolutional network (DeePaN) integrating electronic health records and genomics to discover NSCLC subgroups with differential IO-treatment benefit

Figure 1 illustrates the overall conceptual DeePaN framework. DeePaN employs a graph representation to summarize patient data in an unsupervised autoencoder (AE), hereon referred to as the graph autoencoder (GAE). Specifically, each node in the graph represents a patient with node contents composed of “clinico-genomic” (combined genomic and EHR derived clinical) features; linked neighbor patient nodes share similar clinico-genomic features. The GAE employs a “denoising process” to learn a graph embedding by allowing node content to interact with network features (Methods). The addition of denoising with the GAE is referred as the marginalized graph autoencoder (MGAE)^39^. After application of MGAE based graph embedding, a graph based spectral clustering was then applied to discover patient subgroups with differential IO-treatment benefit.

### DeePaN-identified patient subgroups show distinct IO treatment benefit

Five distinct patient subgroups were identified (Figure 2A) by DeePaN. Overall survival post IO treatments were compared across patient subgroups. The five subgroups showed significant overall survival(OS) differences (log rank test p-value of 2.9 × 10^−9^, median survival ranging from 9.32 to 20.35 months, Figure 2B). This demonstrated DeePaN can effectively discover subgroups with distinct immunotherapy outcomes. Using the overall cohort (1,937 patients) as the control, comparison of survival of each subgroup with the overall cohort identified two subgroups with poor survival, and one subgroup with better survival (Figure 2C). Since the two poor-survival subgroups have similar post-IO OS outcomes (Figure 2B, 2C), we combined them as one single IO non-beneficial subgroup (n=897, 46.3% of the cohort), for comparison to the better survival group as the IO beneficial subgroup (n=400, 20.7% of the cohort). We found significantly different survival post IO between the two groups (log-rank p-value of 2.2 × 10^−11^, median survival of 9.42 vs. 20.35 months, Figure 2D). The demographic and pathologic characteristics of the IO beneficial and non-beneficial subgroups were shown at Supplemental Table 1 in Supplemental material I.

**Figure 2.**
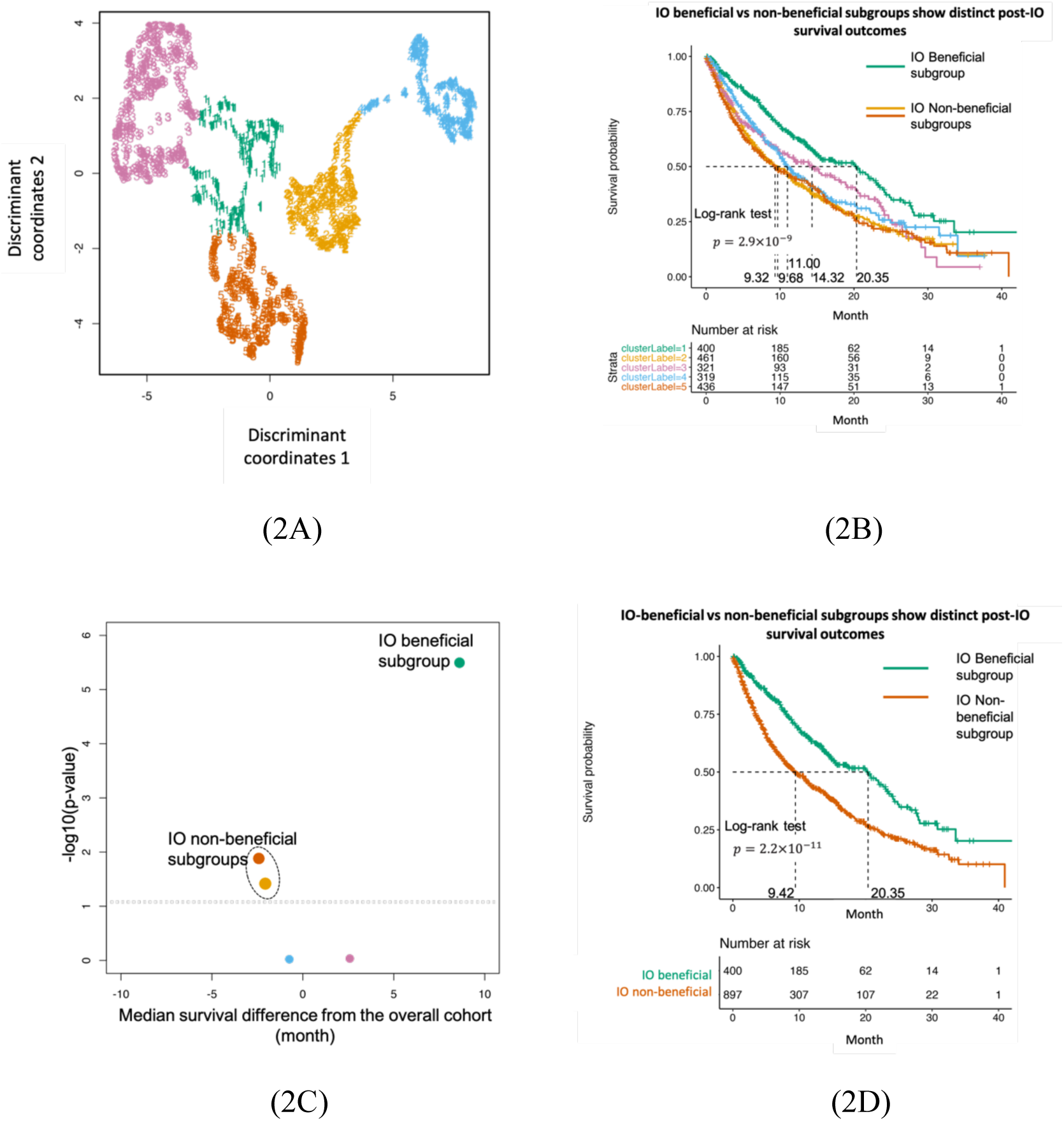
Clinico-genomic “DeePaN” framework discovered NSCLC subgroups with distinct overall survival outcomes of post-IO treatment. **(A)** Five distinct patient subgroups were discovered by DeePaN, visualized by the 2D UMAP projection of the deep patient graph representation in the latent space. Each data point denotes a patient and colors denote distinct subgroup memberships. **(B)** The five subgroups discovered by DeePaN showed significant post-IO overall survival difference by the Kaplan-Meier survival plots (same subgroup color encoding as in A). **(C)** Using the overall cohort (1,937 patients) as the control, comparison of survival of each subgroup with the overall cohort identified distinct IO beneficial and IO non-beneficial subgroups, demonstrated by a volcano plot. Each bubble represents a patient subgroup, same subgroup color encoding was used as in A and B, and bubble sizes are proportional to corresponding subgroup patient counts. The X axis represents the difference of the estimated median survival times between a subgroup and the overall cohort, and the Y axis is the –log10(p-value) of the corresponding log-rank test between a subgroup vs the overall cohort, representing the statistical significance of the observed survival difference. The horizontal dashed line marked the statistical significance cutoff of p-value of 0.05. Two IO non-beneficial subgroups (red and orange) and one IO beneficial subgroup (green) were identified with significantly different post-IO overall survival from the overall cohort. We combined the two IO non-beneficial subgroups (red and orange) into one subgroup since they have similar post-IO survival outcomes. **(D)** The IO beneficial and the combined IO non-beneficial subgroup showed significant (p-value of 2.2 × 10^−11^) post-IO survival difference with estimated median survival of 20.35 months and 9.42 months respectively, by the Kaplan-Meier survival plots.

### Graphical integration of EHR and genomics data is essential towards identifying patient sub-groups with differential IO-treatment benefits

To evaluate whether integration of both EHR and genomics features are essential for effective identification of patient sub-groups with differential IO-treatment benefits, we compared patient grouping using both types of features versus using EHR or genomics features alone. To make a robust comparison, we explored patient grouping with different number of resulting subgroups, including three, five and ten subgroups respectively (Figure 3A). The results demonstrated that integration of both resources was essential to identify patient sub-groups. This highlighted that integration of genomics and real-world clinical phenotype evidence can represent and reveal more of the determinants of cancer patient survival than using genomics or phenotype data alone. Regarding selection of the number of patient subgroups, we selected five subgroups considering both effective subgroup differentiation on IO-treatment benefits and reasonable patient count in each subgroup.

**Figure 3.**
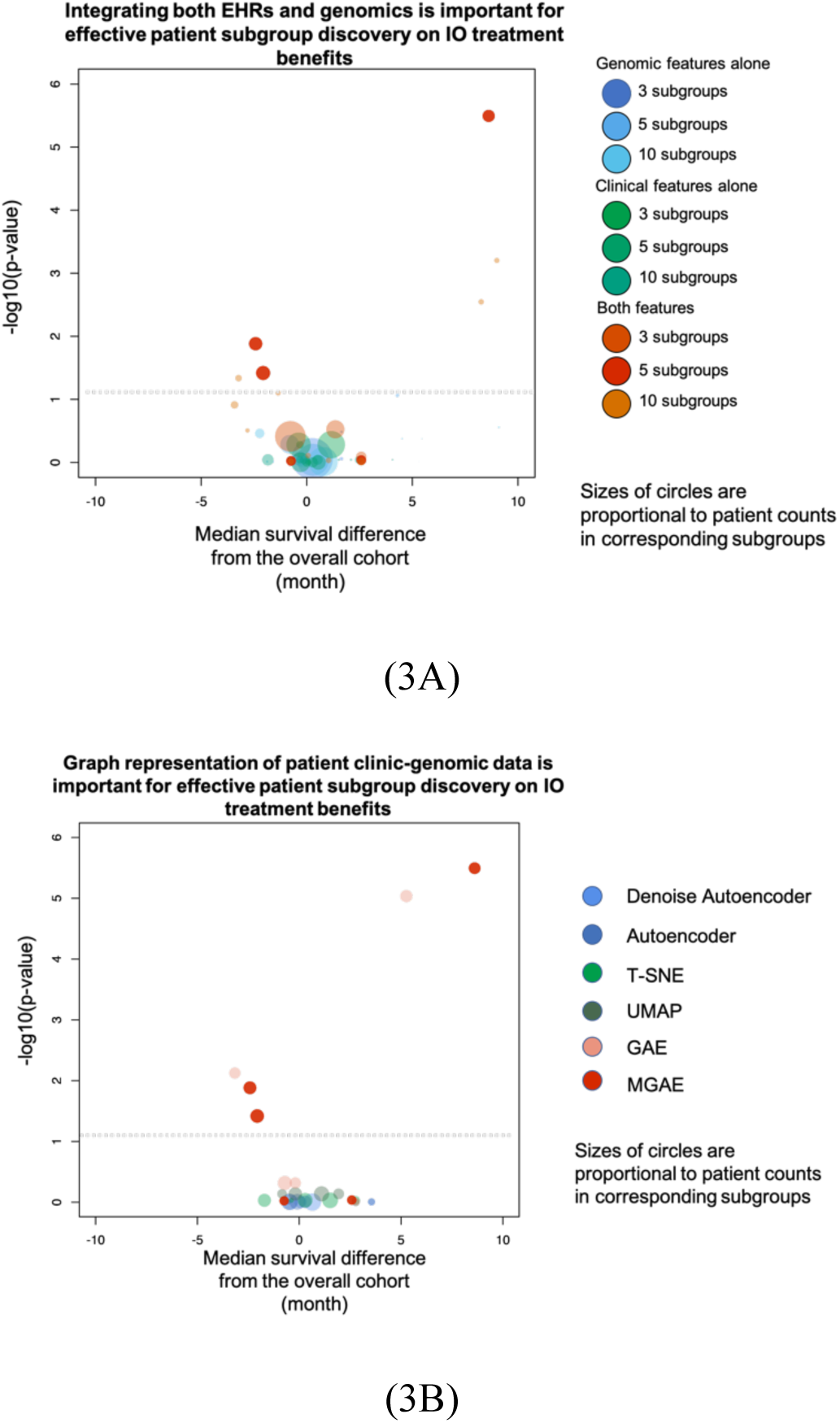
Graph representation of patient data and integration of both EHR and genomics data are essential towards identifying patient subgroups with differential IO-treatment benefits. In both volcano bubble plots, each bubble represents a patient subgroup, the X axis represents the difference of the estimated median survival times between a patient subgroup and the overall cohort as control, and the Y is the –log10(p-value) of the corresponding log-rank test between a patient subgroup vs the overall cohort, representing the statistical significance of the observed survival difference. The horizontal dashed line marked the statistical significance cutoff of p-value of 0.05 (A) Integrating both EHRs and genomics is important for effective patient subgroup discovery on IO treatment benefits. We compared patient subgrouping using both types of features versus using EHR or genomics features alone. To make a robust comparison, we explored different number of resulting subgroups, including three, five and ten subgroups respectively. Integrating both types of features discovers patient subgroups with significantly different post-IO survival, while using EHR or genomic features alone does not identify any subgroups with significantly different post-IO survival. **(B)** Graph representation of patient clinico-genomic data is important for effective patient subgroup discovery on IO treatment benefits. Subgrouping results compared with other methods demonstrates that graph representation of patient data (MGAE and GAE) discovers patient subgroups with significantly different post-IO survival, while non-graph-based approaches (T-SNE, UMAP, autoencoder, and denoise autoencoder) did not identify any subgroups with significantly different post-IO survival.

Additionally, to investigate how 1) the patient-patient relationship based graph topology and 2) denoising process contribute to the effectiveness to stratify patients into subgroups with differential immunotherapy outcomes, we compared four frameworks, our current MGAE which employed both the patient-patient graph topology and the denoising process, 2) the GAE which employed only the patient-patient graph topology but not the denoising process, 3) the denoised Autoencoder which employed only denoising process, and 4) the Autoencoder which employed neither. The results indicate the graph representation of patient-patient relationship is essential since only the MGAE and the GAE are capable to identify sub-groups with differential IO treatment benefits (Figure 3B).Many unsupervised techniques now exist that can accept multi-modal data as input. To further assess the performance of the DeePaN framework we compared it with the commonly used tSNE ^40^ and UMAP ^41^ methods. The results showed that only the DeePaN framework identifies subgroups with differential survival post IO therapy (Figure 3B).

### The clinico-genomic “DeePaN” framework can identify patients with non-high TMB but with beneficial post-IO outcomes

High tumor mutation burden (TMB) is an emerging biomarker utilized to enrich for patients likely to benefit from IO therapy^42,43^, as observed in our flatiron IO cohort (log-rank p-value of 6 × 10^−4^, median survival of 13.3 vs. 24.3 months for TMB non-high vs. TMB high groups respectively, Figure 4A). Many TMB non-high patients, however, may still benefit from IO therapy. We found that subtypes discovered by “DeePaN” were able to further strategy TMB non-high patients into subgroups with significantly differencial survival post-IO therapy (Figure 4B, p-value of 3.8 × 10^−6^ from log-rank test, median survival of 20.8 months and 10.8 months respectively), with about 10 months’ median survival difference between the IO-beneficial vs non-beneficial group. This shows that DeePaN can identify patients with non-high TMB but with beneficial post-IO outcomes with a median survival of over 20 months.

**Figure 4:**
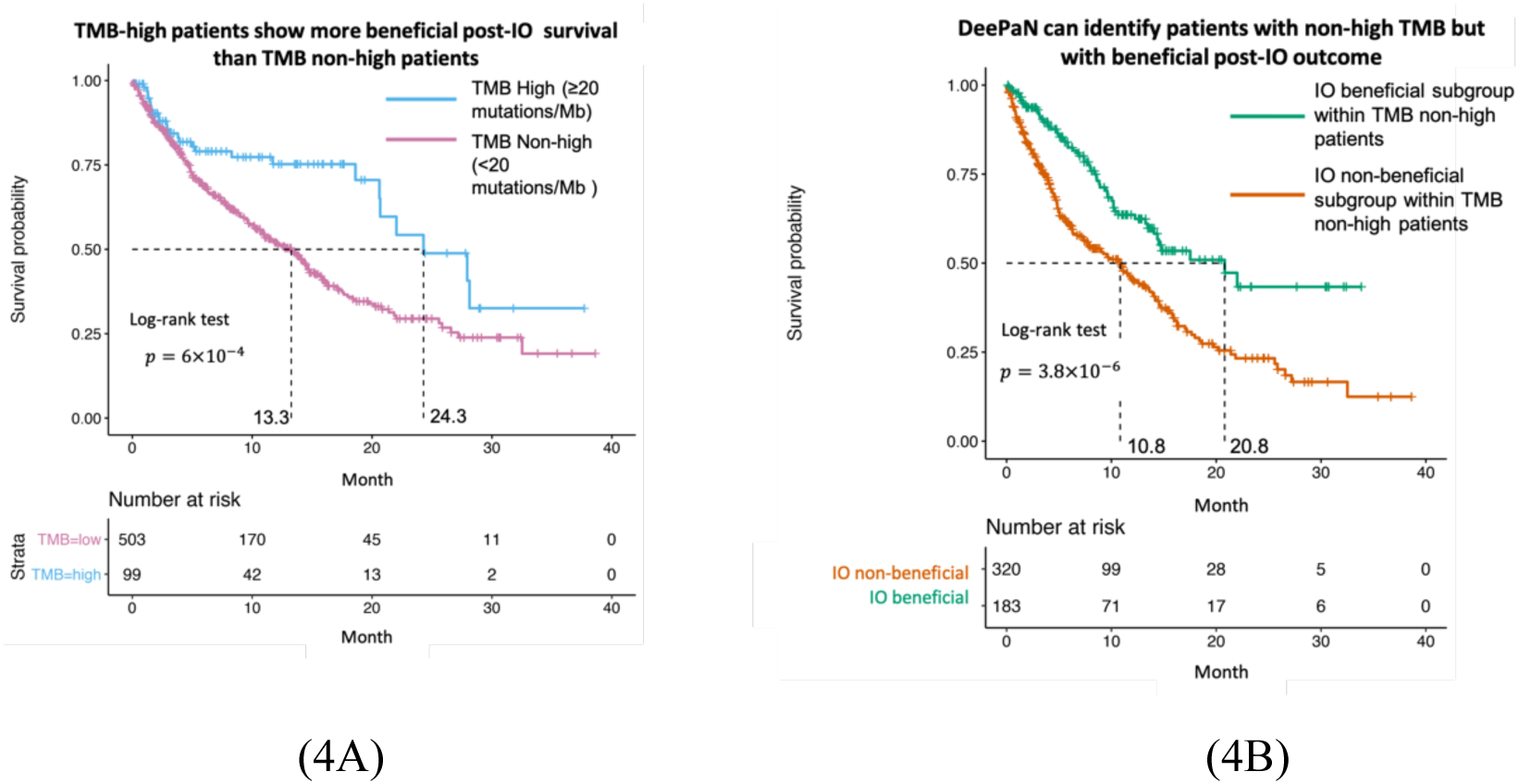
DeePaN can identify patients with non-high TMB but with beneficial post-IO outcomes. (**A**) High tumor mutation burden (TMB) is associated with beneficial post-IO outcomes, as observed in the overall IO cohort (logrank p-value of 6 × 10^−4^, median survival of 13.3 vs. 24.3 months for TMB non-high vs. high groups respectively.) **(B)** DeePaN can identify patients with non-high TMB but with beneficial post-IO outcomes. Subgroups discovered by “DeePaN” are able to strategy TMB-non-high patients into subgroups with significantly differentiated survival post IO therapy (p-value of 3.8 × 10^−6^ from logrank test, median survival of 20.8 months and 10.8 months respectively), with about 10 months’ median survival difference between the IO-beneficial vs non-beneficial group.

### Characterization of the IO beneficial vs non-beneficial subgroups discovered by DeePaN indicates potential to inform therapeutic insight for IO outcome stratification in NSCLC

To inform biological insight of patient stratification with DeePaN, we characterized the IO beneficial vs non-beneficial subgroups identified by DeePaN and identified 21 significantly enriched clinico-genomic features (Supplemental Table 2 in Supplemental material I). Many features have literature evidence indicating relevance to NSCLC prognosis (Supplemental Material II). To explore the potential of DeePaN to real novel and complementary insight in comparison with classical approaches, we further explored the differences in biological insight revealed by DeePaN compared to the classical log-rank test (Supplemental Material II). The log-rank test identified 14 significant features associated with IO outcomes with 8 features in common with DeePaN. 13 out of 21 features enriched between DeePaN defined subgroups did not show a statistically significant relationship to post-IO survival by log-rank, indicating the potential of DeePaN to inform novel insight on IO stratification complementary to the classical approach. For instance, among these 13 features uniquely enriched by DeePaN, features relevant to peripheral immune status such as high blood monocyte count and low blood lymphocyte count are associated with poor post-IO prognosis in NSCLC with supporting literatures ^44-46^; KRAS mutations are enriched with the IO-beneficial subgroup.^47^ There are recent literatures indicating PD-1/PD-L1 blockade monotherapy may be the optimal therapeutic schedule in NSCLC patients harboring KRAS mutations, with KRAS mutations correlating with an inflammatory tumor microenvironment and tumor immunogenicity and thus resulting in superior patient response to PD-1/PD-L1 inhibitors ^47,48^. Taken together, these enriched clinico-genomic features derived from DeePaN discovered subtypes may have potential to inform novel therapeutic insight on IO outcome stratification in NSCLC.

## Discussion

In this study, we explored the feasibility and effectiveness of a graph AI based unsupervised framework, “Deep patient graph” (DeePaN), to stratify IO-treated NSCLC patients from integrating rich genomics and EHR derived clinical data. To the best of our knowledge, our work for the first time has proven the concept that graphical-data-representation based AI can effectively integrate high-dimensional genomic and EHR data to stratify cancer patients relevant to distinct clinical outcomes. This establishes a novel opportunity to use graph AI modeling for precision oncology.

Genomic and EHR data are two major domains of real-world evidence generated in clinical care. Integrative modeling of these data remains challenging but holds great promise to inform precision oncology. Our work demonstrated a graph AI framework can effectively achieve clinico-genomic data integration to inform patient stratification with relevance to outcomes post-IO therapy, and is superior to either data type alone and other stratification methods (Figure 3). For instance, enrichment analysis on patient subgroups identified by DeePaN indicates both clinical features such as blood monocyte count, blood lymphocyte count, and genomic features such as mutated KRAS are potentially associated with differential IO-treatment benefits (supplemental table II in Supplemental material I).

Importantly, the results demonstrate that graph representations of EHR and genomic patient data are important to discover patient sub-groups with differential IO-treatment benefits (Figure 3B). Since our graph representation explicitly constrains patient-patient relationships based on their heterogeneous clinico-genomic feature similarly, the algorithm in turn uses the full heterogeneous feature set when clustering patients rather than over-relying on any one data type. This process is conceptually analogous to clinical diagnosis where a physician relates a patient to a record of similar patients they have seen. Furthermore, leveraging the patient-graph structure, the graph convolutional operation in our framework can iteratively include the information of neighboring nodes for integrating the clinic-genomic data of similar patients. This process propagates to the entire graph, which enables effective patient subtyping from graph clustering at the global scale.

Characterization of the IO beneficial vs non-beneficial subgroups identified by DeePaN indicates potential to inform novel and complementary therapeutic insight for IO stratification in NSCLC in comparison with classical approaches such as the logrank test approach (Supplemental material II). Mechanistic insight on IO outcomes in NSCLC was indicated by features significantly enriched by DeePaN discovered patient subgroups but not reaching statistical significance by logrank test. For instance, the enrichment of high blood monocyte count and low lymphocyte count in the IO non-beneficial group identified by DeePaN indicates host peripheral immune status may contribute to IO outcomes; the enrichment of mutated KRAS in the IO beneficial subgroup was supported by literature evidence that KRAS mutations correlating with an inflammatory tumor microenvironment and tumor immunogenicity and thus resulting in superior patient response to PD-1/PD-L1 inhibitors in NSCLC.^47^ Another DeePaN unique finding is the enrichment of mutated NKX2.1 gene in the IO-beneficial subgroup. NKX2.1 is a proto-oncogene contributing to lung cancer development, literature evidences are debating the role NKX2.1 in lung cancer prognosis, our finding supports to continue to explore its role on post-IO prognosis.^49^

There are opportunities for future work. First, in EHRs, the existence of an assay result or the design of the treatment plan for a patient can be the result of comprehensive factors including economic stabilities, educations, community and social context, et al. One aspect of future work is to include more features such as social economic conditions et al to into modeling. Second, as a graph based AI framework, DeePaN utilized both the non-linear combination of clinico-genomic features and the patient graph structure for effective subtype identification, it remains challenging to biologically interpret this process^50^. We utilized enriched clinico-genomic features derived from DeePaN discovered patient subtypes to inform therapeutic insight, which can be improved by future work of developing more interpretable graph-AI models such as graph attention networks^51^ to understand what drives the patient stratification to inform biomarker and therapeutic insight discovery. To validate “DeePaN” discovered patients’ subtypes to inform clinical insight, we suggest that, as many researchers have argued^50,52,53^ and the U.S. Food and Drug Administration has been advocating^54,55^ and practicing^56^, artificial intelligence models should be considered as medical devices or drugs and thus the effectiveness and safety should be evaluated through randomized clinical trials, including EHR-based pragmatic trials. A future direction will be to use multi-site randomized pragmatic trials to examine the effectiveness of the identified subtypes in augmenting clinical decisions on immunotherapies.

Our work thus provides the first evidence that integrative modeling using genomics and EHR data in a graph AI framework has clinical utility in precision oncology. As a case study, we show that as an emerging IO biomarker, although TMB-high vs TMB-non-high groups are associated better and worse post-IO outcomes respectively, the TMB-non-high group may contain heterogeneous patient population with distinct post-IO outcomes (Figure 4A, 4B). Importantly, patient subgrouping discovered from our DeePaN framework can effectively stratify the heterogeneous TMB-non-high group to identify patient subtypes with non-high TMB but beneficial IO outcomes. This highlights the potential clinical utility of our framework on augmentation of the TMB IO biomarker. Characterization of the IO beneficial vs non-beneficial subgroups discovered by DeePaN indicates potential to inform therapeutic insight to stratify NSCLC patients on IO outcomes. The “deep patient graph convolutional network” approach can be potentially applied in a wide range of clinical applications. For example, by incorporating other types of treatment regimens such as targeted therapies, chemotherapies, radiotherapies et al, this methodology can be used for recommending therapies for NSCLC patients. Similarly, this approach can be applied in other cancer types or non-cancer diseases to inform precision medicine. Besides unsupervised subtyping, representation of the original clinic-genomic data in latent space from a graph embedding can also be used for supervised learning to predict disease diagnosis or prognosis, for health trajectory projection, and so on. Our approach thus paves new ways in effectively using clinico-genomic graph AI modeling for diverse applications in precision medicine.

In summary, our work serves as a proof-of-concept study to demonstrate that a patient-graph based AI framework is feasible and effective to integrate EHR and genomic data to inform precision oncology. With the continuous advancement of various graph-building tools and graph AI methods, we will expand our work to incorporate them to continue to inform more precision-medicine questions.

## Materials and Methods

### Study Design

The aim of this study is to explore the feasibility and effectiveness of a data-driven, graph AI based unsupervised framework to strategy IO-treated NSCLC patients into subgroups with distinct immunotherapy outcomes by integrating rich genomics and EHR data. To define immunotherapy outcomes, we focused on the overall survival of the NSCLC population since the start date of the first IO treatment. The clinical and genomic features were defined as baseline features measured before the start of the IO therapies.

This is a secondary analysis of pre-existing, de-identified, retrospective electronic medical record data and therefore IRB review is not required.

### Patient cohort and endpoint

The NSCLC IO study cohort and dataset were established from the Flatiron Health longitudinal EHR-derived database including RWE genomics and clinical data curated from the EHR data of over 270 cancer clinics representing more than 2 million active patients across the United States. The Foundation Medicine genomic testing data in this database was from January 2010 to October 2018. The inclusion criteria of the cohort were (See Supplemental Figure 1A in Supplemental material I): NSCLC patients identified with International Classification of Diseases (ICD) code for lung cancer (ICD-9: 162.x; ICD-10: C34.x or C39.9)^38^, evidence of administration of checkpoint inhibitors anti-PD-1/PD-L1 agents either as monotherapy or as part of a combination regimen^38^, and with the Foundation Medicine genomic testing data available.

The endpoint is defined as the overall survival of post-IO treatment. The overall survival time was defined as the length of time from the first use of IO therapies to the event of deceased patients, or to last follow-up date.^38^

### Clinical features and genomic features

The clinical and genomic features were defined as baseline features measured within 6 months before the start of the IO therapies. Clinical and genomic features were screened according to prior knowledge and data availability. Totally 52 clinical features and 48 genomics features were used in our work.

Clinical features included: 1) demographics: race, gender; 2) behavioral: smoking status; 3) vitals: body weight, body height, oxygen saturation in arterial blood by pulse oximetry; 4) medical history: lines of IO therapy; 5) pathological features: eastern cooperative oncology group (ECOG) performance status, cancer stage, 6) pathological staining of biomarkers: ALK, BRAF, EGFR, KRAS, ROS1, PDL1 in tumor cells, and PDL1 in tumor infiltrated lymphocytes (TIL); 7) laboratory measurements available in more than 800 patients: leukocytes, hemoglobin, platelets, hematocrit, erythrocytes, serum creatinine, urea nitrogen, alanine aminotransferase, serum sodium, serum potassium, aspartate aminotransferase, alkaline phosphatase, serum albumin, bilirubin, serum protein, lymphocytes per 100 leukocytes, calcium, lymphocytes, monocytes per 100 leukocytes, serum glucose, serum chloride, monocytes, neutrophils, basophils per 100 leukocytes, glomerular filtration rate, basophils, eosinophils per 100 leukocytes, eosinophils, serum magnesium, granulocytes per 100 leukocytes, neutrophils, lactate dehydrogenase, and serum ferritin. (See Supplemental material I, Supplemental Figure 1B is the visualization of clinical features); 8) Foundation Medicine derived features: PDL1 expression levels in tumor cells, PDL1 expression levels in TIL, tumor mutation burden (TMB)^38^ (High if TMB >= 20 mutations/MB; non-high if TMB < 20 mutations/MB)^38^, and microsatellite instability (MSI).

Genomic features are based on tumor sequencing of FoundationOne platform, which includes full exonic coverage of 395 genes and intronic analysis for rearrangements at a depth of 500-1000x^38^. Genomic features include known and likely genomic alterations occurring in at least 50 patients at the gene level, including the following genes (sorted by frequency, see Supplemental Figure 1C in Supplemental material I): “TP53”, “KRAS”, “CDKN2A”, “STK11”, “CDKN2B”, “EGFR”, “PIK3CA”, “LRP1B”, “MYC”, “KEAP1”,“NF1”, “NKX2.1”, “PTEN”, “SMARCA4”, “ARID1A”, “RBM10”, “RB1”, “SOX2”,“NFKBIA”, “CCND1”, “FGF3”, “FGF4”, “FGF19”, “BRAF”, “MLL2”, “ATM”,“MDM2”, “ERBB2”, “TERC”, “MET”, “SPTA1”, “FGFR1”, “RICTOR”, “MCL1”,“DNMT3A”, “ARID2”, “PRKCI”, “FAT1”, “ZNF703”, “TERT”, “APC”, “NFE2L2”,“FGF12”, “MYST3”, “FRS2”, “TET2”, “PTPRD”, and “CCNE1”.

### Problem formulation

Given the NSCLC patient data with clinico-genomic features, we formulate the task of patient subgrouping as a graph clustering problem on an undirected graph encoding patient-patient relationships. Specifically, patients are represented as nodes in the graph, and patients with similar clinico-genomic features are linked by edges.

It is beneficial to formulate the patient-patient relationship into a graph since both the node content (patient clinical and genomic features) and structure interaction (patient-patient connectivity based on feature similarity) will be used and integrated. We model the patient-patient relationship using a graph *G* with node content as *G* = (*V, E, X*) with *N* nodes (patients) *V*_*i*_∈ *V, i* ∈ [0, *N*], edges connectivity (*V*_*I*_,*V*_’_) ∈ *E*, where the edge connectivity can be either 0 (disconnected) or 1 (connected), and *x*_*i*_ ∈ *X, i* ∈ [0, *N*] is the attribute vector associated with vertex *V*_*i*_. Each patient has an attribute vector of clinico-genomic features of length *d* (e.g. age; gender; LDH lactate dehydrogenase measurement, gene mutation status et al) as node features. The patients’ clinico-genomic features are encoded by categorical vectors *X* (see Supplemental material I for details) and two nodes (patients) are linked if node feature vectors are similar (see Supplemental Material I for details). So formally, the graph can be represented by two types of information, the patient content information *X* ∈ *R*^*n*×*d*^ and the structure information *A* ∈ *R*^*n*×*n*^, where A is an adjacent matrix of *G* and *A*_*I,j*_= 1 if *e*_*i,j*_∈ *E* otherwise, 0.

Given a patient-patient graph *G*, patient subtyping is then to partition the patient nodes from *G* into *k* disjoint subgroups {*G*_1_, *G*_2_, …, *G*_*k*_} so that different patient subgroups may have differentiable clinical outcomes (here we use overall survival post-IO treatment).

### Implementation

To achieve above-mentioned goal, we need to solve two main tasks: 1) to learn informative patient feature representation for downstream graph clustering method to work properly; 2) to discover new patient clusters (subgroups) on the graph that have differentiable clinical meaning.

#### 1) Learn patient deep feature graph representation

To fully extract and have deep feature representation, we apply the marginalized graph autoencoder (MGAE) method ^39^ to exploit the patient network information. The MGAE is based on graph convolutional network (GCN) ^36^ and to learn the convolution feature representation on the structure information with node content in the spectral domain. MGAE can exploit the interplay between node content and graph structure information by using a marginalization process, which is to encode content features of the graph into the deep learning framework. The reconstructed feature representation can be achieved by training an MGAE^39^ on this patient network using the objective function as following:

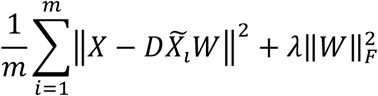

where X = {*x*_*1*_, …, *x*_*n*_} ∈ *R*^*n*×*d*^ is the input, *D* is the degree matrix, *W* is trainable weights, and *m* is the number of corruption times. (Please refer to Methods in Supplemental Material I for more details)

To learn a deep feature representation of patients’ network, we built up the network in a deep layer fashion by stacking multiple layers of autoencoders (Figure 1). The patients’ representation from the output (*l* − 1)-th layer *z*^(*l*-1)^ can be then used as input of the *l*-th layer. Our framework was constructed in three layers; more layers for a graph network are unnecessary since all neighboring information can be fully explored and integrated after three layers for a specific node (See experiments of exploration number of layers for a graph network in Kipf *et al*^36^). We used the reconstructed output from the last layer as the high-level patients’ representation for downstream analysis, i.e. detection of new patient subgroups.

#### 2) Discovery of patient subgroups

The learned representation *z*_0_ for the patients’ graph, which is reconstructed from MGAE’s representation (integration of both content and structure information), can then be used to discover patient subgroups. Before directly applying spectral clustering, we refine the reconstructed representation *Z*_0_ as following:

1. apply a linear kernel function to achieve 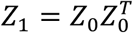 to learn the pairwise relationship for the patient node.
2. ensure the representation is symmetric and nonnegative, we achieved normalized Laplacian 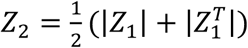

New clusters (i.e. patient subgroups) were identified using a spectral clustering algorithm, which was done by running k-means on the top number of clusters eigenvectors of the normalized Laplacian *Z*_2_. Those clusters are identified as new patient subgroups. We used the Kaplan-Meier (KM) estimate^57^ to assess if discovered subgroups have differentiable post-IO survival outcomes to inform patient stratification benefiting from IO therapies.

## Data Availability

The data source is from the Flatiron NSCLC clinico-genomic Database. Since the data resource is patient data from EHRs, it is not available for public share.

## Data availability

Our implementation is based upon MGAE’s open-source code: https://github.com/FakeTibbers/MGAE.

## Acknowledgements

This research was supported by AstraZeneca (AZ) Postdoc Funding. The high-performance computing resource was supported by Scientific Computing Platform (SCP) at AZ. D.X.’s work was partially supported by the National Institutes of Health (R35-GM126985). J.S.’s work was partially supported by the Cancer Center Support Grant from the National Cancer Institute to the Comprehensive Cancer Center of Wake Forest Baptist Medical Center (P30 CA012197). We would like to thank our colleagues Kris Sachsenmeier, Gayle Pouliot, Zhongwu Lai, Melinda Merchant, Mingchao Xie, Krishna Bulusu, and Steven Criscione for their helpful discussion and constructive suggestions.

## Authors’ contributions

B.L. J.D., and J.S. designed and directed this study. D.X. provided scientific suggestions and helpful discussion. C.F. collected and processed the data, built the model, trained the model, carried out experiments. All authors analyzed and validated the experimental results of data analysis. All authors wrote, reviewed and revised the manuscript.

## Additional information

**Supplementary Information** accompanies this paper available online.

